# Exploration of the potential of high resolution and contrast 7 Tesla MR brain imaging in neonates

**DOI:** 10.1101/2023.09.28.23296232

**Authors:** Philippa Bridgen, Raphael Tomi-Tricott, Alena Uus, Daniel Cromb, Megan Quirke, Jennifer Almalbis, Beya Bonse, Miguel De la Fuente Botella, Alessandra Maggioni, Pierluigi Di Cio, Paul Cawley, Chiara Casella, Ayse Sila Dokumaci, Alice R Thomson, Jucha Willers Moore, Devi Bridglal, Joao Saravia, Thomas Finck, Anthony N Price, Elisabeth Pickles, Lucilio Cordero-Grande, Alexia Egloff, Jonathan O’Muircheartaigh, Serena J Counsell, Sharon L Giles, Maria Deprez, Enrico De Vita, Mary A Rutherford, A David Edwards, Joseph V Hajnal, Shaihan J Malik, Tomoki Arichi

**Author notes:** Joint senior authors (Shaihan J Malik, Tomoki Arichi). Corresponding author: Dr Tomoki Arichi Department of Perinatal Imaging, King’s College London 1^st^ Floor South Wing, St Thomas’ Hospital, Westminster Bridge Road, London, SE1 7EH, UK.

## Abstract

**BACKGROUND:** Ultra-high field MR imaging offers marked gains in signal-to-noise ratio, spatial resolution, and contrast which translate to improved sensitivity for pathology and anatomy. These benefits are particularly relevant for the neonatal brain, as it is rapidly developing and sensitive to injury. However, experience of imaging neonates at 7T has been limited due to regulatory, safety, and practical considerations.

**PURPOSE:** To establish a program for safely acquiring high resolution and contrast brain images from neonates on a 7T system.

**STUDY TYPE:** Prospective case series.

**POPULATION:** Images were safely acquired from 35 neonates on 44 occasions (median age 39+6 postmenstrual weeks, range 33+4 to 52+6; median body weight 2.93kg, range 1.57to 5.3kg) over a median of 49 mins 30 sec.

**FIELD STRENGTH/SEQUENCE:** 7T, acquired sequences included T2 weighted (TSE), Actual Flip angle Imaging, functional MRI (BOLD EPI), susceptibility weighted imaging, and MR spectroscopy (STEAM).

**ASSESSMENT:** Peripheral body temperature, physiological measures (heart rate, oxygen saturations). Review of acquired images by Neonatal Neuroradiologists for visual identification of anatomy and pathology, and by radiographer and researcher for assessment of image quality.

**STATISTICAL TESTS:** Two tailed paired t-test, P<0.05 was considered statistically significant.

**RESULTS:** There was no significant difference between temperature before and after scanning (p=0.76). Image quality assessment compared favourably to state-of-the-art 3T acquisitions. Anatomical imaging demonstrated excellent sensitivity to structures which are typically hard to visualise at lower field strengths including the hippocampus, cerebellum, and vasculature. The potential of 7T imaging is highlighted using contrast mechanisms which are enhanced at ultra-high field including susceptibility weighted imaging, functional MRI, and MR spectroscopy.

**DATA CONCLUSION:** We demonstrate safety and feasibility of imaging vulnerable neonates at ultra-high field. Our preliminary imaging suggests ultra-high field has untapped potential to provide important new insights into brain development and pathological processes during this critical phase of early life.

## INTRODUCTION

During the neonatal period (the time of birth and the first few weeks of postnatal life), the human body undergoes a series of highly programmed yet rapid sequences of physiological and anatomical maturation which enable adaption to the demands of the new ex utero environment. In this critical juncture, dramatic developmental changes are similarly seen within the human brain, as the cortex rapidly expands and folds and axonal projections proliferate within the white matter ^1^, neurotransmitter action and receptor density evolve ^2^, and distinct correlated network patterns of functional activity emerge ^3^. The fundamental importance of this period is emphasized by the effects of congenital or acquired perinatal brain pathology, which invariably result in life-long alterations in brain structure and function and can lead to later neurodisability. Furthermore, multiple lines of evidence now suggest that the previously unknown underpinnings of the brain abnormalities that underlie mental health disorders and neurological diseases in adulthood also likely begin in the perinatal period, well before symptoms or behavioural difficulties manifest later in life ^4^. There is therefore a clear need for tools which can not only accurately identify pathology in the neonatal period but can also act as novel biomarkers for later outcome and provide mechanistic insight into pathophysiological processes to inform potential treatments.

In recent years, MR imaging at ultra-high field (7 Tesla (T) or higher) has become increasingly used in adult subjects due to the significant gains in signal-to-noise-ratio (SNR) and differences in tissue contrast it offers over standard MR systems (1.5 or 3T) ^5^. This has proved to be particularly clinically beneficial for enhancing diagnosis and understanding about diseases which affect the cortex such as drug-resistant focal epilepsy where no relevant lesion has been identified at standard field strengths ^6^ and cerebral vasculature such as small-vessel stroke ^7^. Furthermore, numerous studies have demonstrated that ultra-high field imaging can provide profound new insights into brain physiology, particularly using contrast mechanisms based on magnetic susceptibility such as blood-oxygen-level-dependent (BOLD) functional MRI (fMRI), where the increased sensitivity and spatial specificity enable characterization of neural activity at a cortical laminar level ^8^; and Magnetic Resonance Spectroscopy (MRS) where there is improved separation of overlapping frequency peaks ^9^. Imaging at 7T for neonates is therefore a compelling prospect, as it could theoretically enable more detailed visualization of key developing brain structures, improved understanding about pathophysiological processes, and reduced prognostic uncertainty.

Despite the likely benefits, regulatory barriers and safety concerns have thus far prevented widespread application of 7T MR imaging in neonates, with only a single study previously demonstrating feasibility ^10^. Furthermore, detailed safety modelling has demonstrated that careful planning is required as there are higher risks associated with increased specific absorption rate (SAR) and temperature instability in neonates compared to adults under the same conditions at 7T ^11^. A further consideration is that the composition of neonatal brain tissue differs markedly from that of the mature adult brain, and thus image acquisition sequence parameters must be adapted accordingly to account for significantly different field-dependent tissue relaxation times ^12^. With these factors in mind, we aimed to establish a 7T imaging program for neonates following a comprehensive safety assessment. We describe our resulting safety approach, initial experiences, and results, and demonstrate the potential gains in anatomical and pathological sensitivity for the neonatal population.

## MATERIALS AND METHODS

All examinations took place on a Siemens Magnetom Terra 7T system (Siemens Healthineers, Erlangen, DE) in the London Collaborative Ultra-high field System (LoCUS) centre at St Thomas’ Hospital London. National ethics committee approval (NHS REC: 19/LO/1384) was attained for the work and all images were acquired following parental consent.

### Safety

Previous neonatal modelling suggests that close monitoring of infant body temperature is imperative for MRI scanning, as neonates can become rapidly hypothermic if not sufficiently insulated (due to reduced subcutaneous fat in comparison to adults) ^13^ or could become hyperthermic due to systemic heating inherent in prolonged radiofrequency (RF) exposure ^11, 14^. The latter is an important consideration at ultra-high field as in addition to their small body size, the higher water content of neonatal tissue leads to significantly different dielectric properties in comparison to adults. Taking this into account, a recent detailed RF safety simulation using a head-centered model within a local transmit coil of the generic type deployed in the Siemens Terra scanner, has shown that SAR is likely elevated in neonates at ultra-high field in comparison to adults being scanned under the same conditions ^11^. This is in contrast to lower field strengths (3 Tesla or less) where heating was predicted to be less. As the study for 7T indicated that systemic (rather than localized) heating posed the most likely risk, further mitigation was achieved by mandating continuous monitoring of axillary temperature in addition to heart rate and oxygen saturation throughout the examination. This was achieved using a Philips Invivo Expression MR400 monitor (Philips Healthcare, Gainesville, FL) which was tested to verify safety/efficacy for use at 7T ^15^. To further mitigate the risk of potential temperature effects and following an externally led local risk assessment, the scanner software was modified for imaging neonates to use a more conservative SAR estimation, with estimated SAR increased by a factor of 2.8 for equivalent power. A robust process was developed to allow this to occur safely: software changes result in the displayed name of the coil changing (as a visual check) and then updated SAR estimates are experimentally confirmed by scanning a phantom before and after switching. The process is set out using a detailed checklist which must be signed-off before each scanning session by two authorized personnel.

### Image Acquisition

Images were successfully acquired from 35 neonates (24 male) of median postmenstrual age at scan: 40+0 weeks (range 33+4 to 52+6 weeks); median gestational age at birth: 35 weeks (range 27+6 to 42+1 weeks); median body weight at scan: 2.9kg (range 1.6 to 5.3kg). Examinations took place on 44 occasions lasting a median total of 49 min 30 sec (range 20-77 minutes), with 41min 15sec of active scanning when the full core protocol was completed. 9 infants were imaged both in the preterm period and at term equivalent age; and one infant with congenital heart disease was imaged both pre-operatively and post-operatively. 18 infants additionally had images acquired on a 3T system (Achieva, Philips, Best NL) with 32ch receive head coil, with a further subset of 5 infants having the 3T scan within 24 hours of the 7T scanning session for direct comparison. All Images were reviewed by experienced neonatal neuroradiologists (AE, JS, TF, MAR).

Participants were positioned supine headfirst in the isocenter of a 1TX-32RX Nova head coil with the aid of foam and inflatable pads (Pearltec, Zurich CH) to help reduce head movement and to ensure that maximum signal was received from all elements of the head coil. Participants were scanned during natural sleep following feeding and were then swaddled and immobilized inside 2 pre-warmed blankets and a vacuum evacuated bag (Pearltec, Zurich CH). Hearing protection was applied using molded dental putty in the external auditory meatus (President Putty, Coltene Whaledent, Mahwah, NJ, USA). Two infants with congenital cardiac disease were receiving a continuous intravenous prostaglandin E1 (PGE1) infusion (with the infusion pumps outside the examination room) during image acquisition to maintain patency of the ductus arteriosus. All scans were supervised by experienced clinical staff (a doctor and nurse) with appropriate training in neonatal care and resuscitation who also reviewed the neonates’ temperature and vital sign measurements throughout scanning.

In all infants, the following were acquired high-resolution 2D T2-weighted images (T2W) in at least 2 orthogonal planes, mid brain and cerebrum, MR spectroscopy (MRS) including metabolite T1 estimation, 3D susceptibility weighted imaging (SWI), and Blood Oxygenation Level Dependent (BOLD) functional MRI (fMRI) were acquired. Actual Flip angle Imaging (AFI) B_1_ ^+^ mapping ^16^ was obtained in 15/44 cases (due to time constraints). Sequence parameters were selected initially from those optimized at 3T for the developing Human Connectome Project ^17^ and were then iteratively adapted to maximize SNR and resolution (*parameters for all acquisitions are detailed in Table 1*).

**Table 1:** Acquisition sequence parameters. (FOV: field of view; TR: repetition time; TE: echo time; FA: flip angle; TI: inversion time; GRAPPA: GeneRalized Autocalibrating Partial Parallel Acquisition; TM: mixing time; STEAM: STimulated Echo Acquisition Mode; AFI: Actual Flip-angle Imaging; GRE-EPI: Gradient echo Echo Planar Imaging)

| Sequence | Resolution (mm) | FOV Frequency | TR (ms) | TE (ms) | Acceleration factor | FA (deg) | Other Parameters | Acquisition Time (min) |
| --- | --- | --- | --- | --- | --- | --- | --- | --- |
| Localizer | 0.7x0.7x5.0 | 250 x 250mm | 4000 | 107 | GRAPPA 4 | 115 |  | 0:22 |
| B0 map | 5.0x5.0x5.0 | 210 x220mm | 10.0 | 1.02, 2.26, 4.08 | none | 10 |  | 0:19 |
| T2 - Axial | 0.6x0.6x1.2 | 141 x 151mm | 8640 | 156 | GRAPPA 2 | 120 | - | 2:37 |
| T2 – Sagittal | 0.6x0.6x1.2 | 154 x 125mm | 8640 | 156 | GRAPPA 2 | 120 | - | 3:12 |
| T2 – Coronal | 0.6x0.6x1.2 | 154 x 125mm | 8640 | 156 | GRAPPA 2 | 120 | - | 3:29 |
| AFI | 2.2x2.2x3.0 | 140 x 140 | 141 | 1.93, 3.62, 9.00 | GRAPPA 3 | 60 |  | 1:54 |
| MRS (STEAM) | 16x16x16 | - | 3000 | 20 | none | 90 | TM = 10ms | 2:45 |
| BOLD fMRI (GRE-EPI) | 1.0x1.0x1.0 | 125 x 125mm | 2560 | 43 | GRAPPA 3 Multiband 3 | 90 (exc) 110 (deg) | - | 7:26 |
| Susceptibility Weighted Imaging (SWI) | 0.2x0.2x1.2 | 122 x 150mm | 22 | 15 | GRAPPA 3 | 15 | - | 2:12 |

To address intra-volume motion artifacts, as well as provide increased signal-to-noise ratio (SNR) and image resolution, T2 images of different orientations were combined into a single high-resolution volume with isotropic resolution 0.45mm using Slice to Volume Reconstruction (SVR) as implemented in SVRTK (https://svrtk.github.io). Each acquired image was split into odd and even stacks of slices, and heavily motion corrupted resulting stacks were excluded after visual inspection. Remaining stacks were co-aligned using volumetric registration and reconstructed into a single isotropic volume using a super-resolution algorithm ^18^. Reconstructed images were then tissue segmented, region parcellated and surfaces generated using the developing Human Connectome Project (dHCP) structural pipeline (https://github.com/BioMedIA/dhcp-structural-pipeline) ^19^. Native and SVR image quality were assessed by visual review of the T2-weighted images with two reviewers (PB, TA) using a grading system of 1-4 previously used in the dHCP ^19^: where 1 is a poor quality image, 2 is an image with significant motion artifact, 3 is an image with negligible motion artifact; and 4 is a good quality image (*example images shown in Figure 1a*).

**Figure 1:**
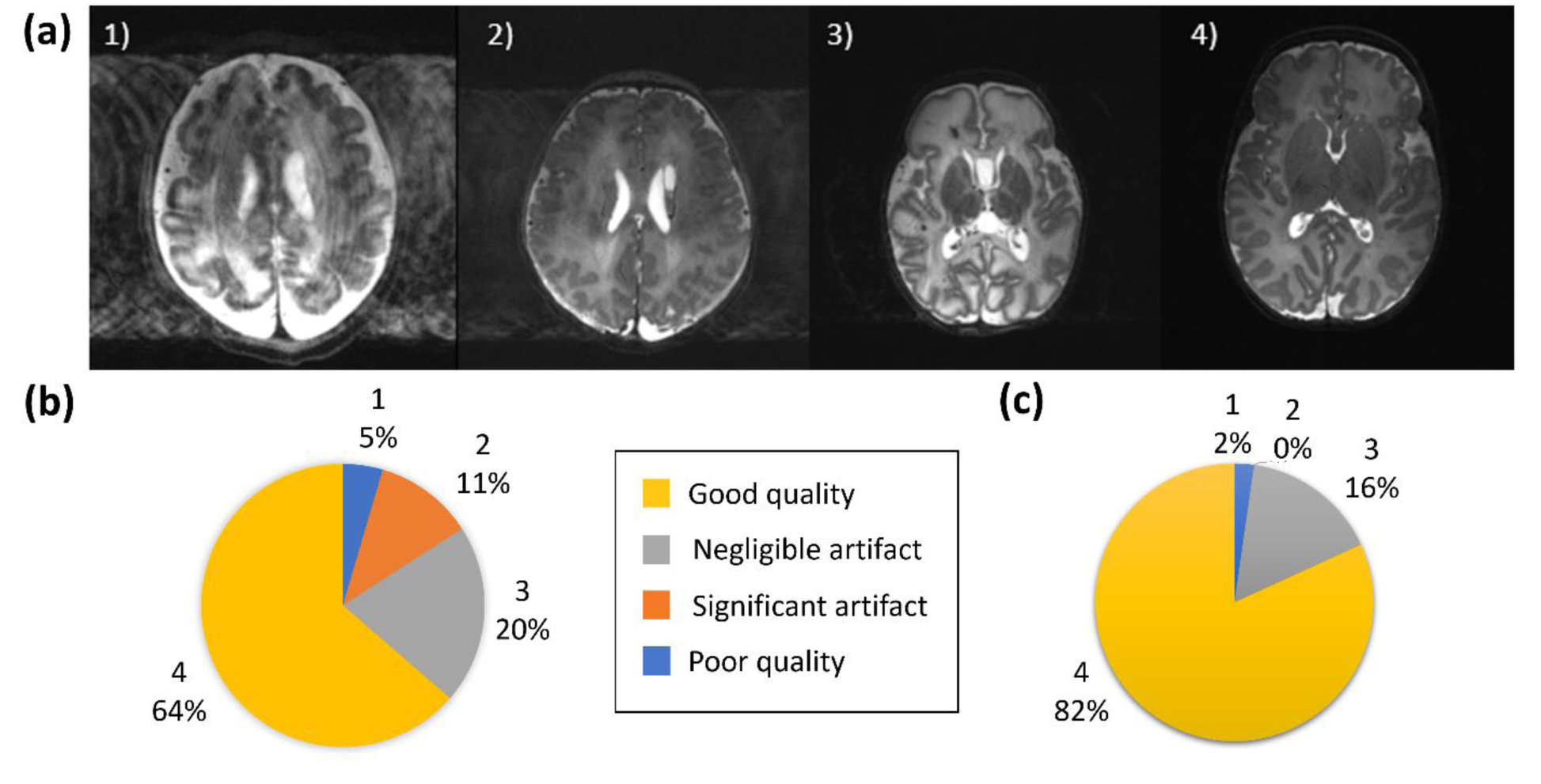
Image quality assessment. (a) Shown are representative images depicting quality of the native images as assessed using a previously described score ^19^, where 1) is a poor quality image 2) image contains significant motion artefact, 3) image with negligible motion artefact and 4) good quality image. (b) Image quality assessment results for the native T2-weighted images and (c) for the slice-to-volume reconstructed T2-weighted images.

BOLD fMRI resting state networks were delineated following standard preprocessing steps including rigid body motion correction, high pass temporal filtering, slice time correction, and spatial smoothing (Gaussian filter of full-width half maximum 3mm) and using independent component analysis (ICA) as implemented in MELODIC v3.0, part of the FSL software library (www.fmrib.ox.ac.uk/fsl) ^20^. Minimum intensity projection (MIP) images were prepared from the magnitude and phase SWI data using CLEAR-SWI ^21^. Metabolite concentrations were estimated from the MRS data following pre-processing using Suspect-MRS (https://suspect.readthedocs.io/en/latest/) including optimal channel-combination ^22^, eddy current correction, spectral registration and outlier rejection ^23^. LCModel ^24^ was then used for metabolite fitting with basis sets generated by Tarquin (brain+Gly+Glth) ^25^.

## RESULTS

All infants tolerated scanning on the 7T system, with stable vital signs throughout the scan session. There were no adverse events during image acquisition and there were no concerns expressed about discomfort or altered behaviour reported by clinical staff or parents either during or in the 24 hours subsequent to the scanning session. Infant axillary temperature remained stable throughout image acquisition, with no significant difference in temperature measurements at the start of scanning (median 36.9 degrees centigrade, range 35.5-37.4) and at the end of scanning (median 36.8 degrees centigrade, range 35.6- 37.7) on the 7T system (paired t-test: p=0.76, *Figure 2*). This included the youngest preterm infant studied, who weighed 1.58kg.

**Figure 2:**
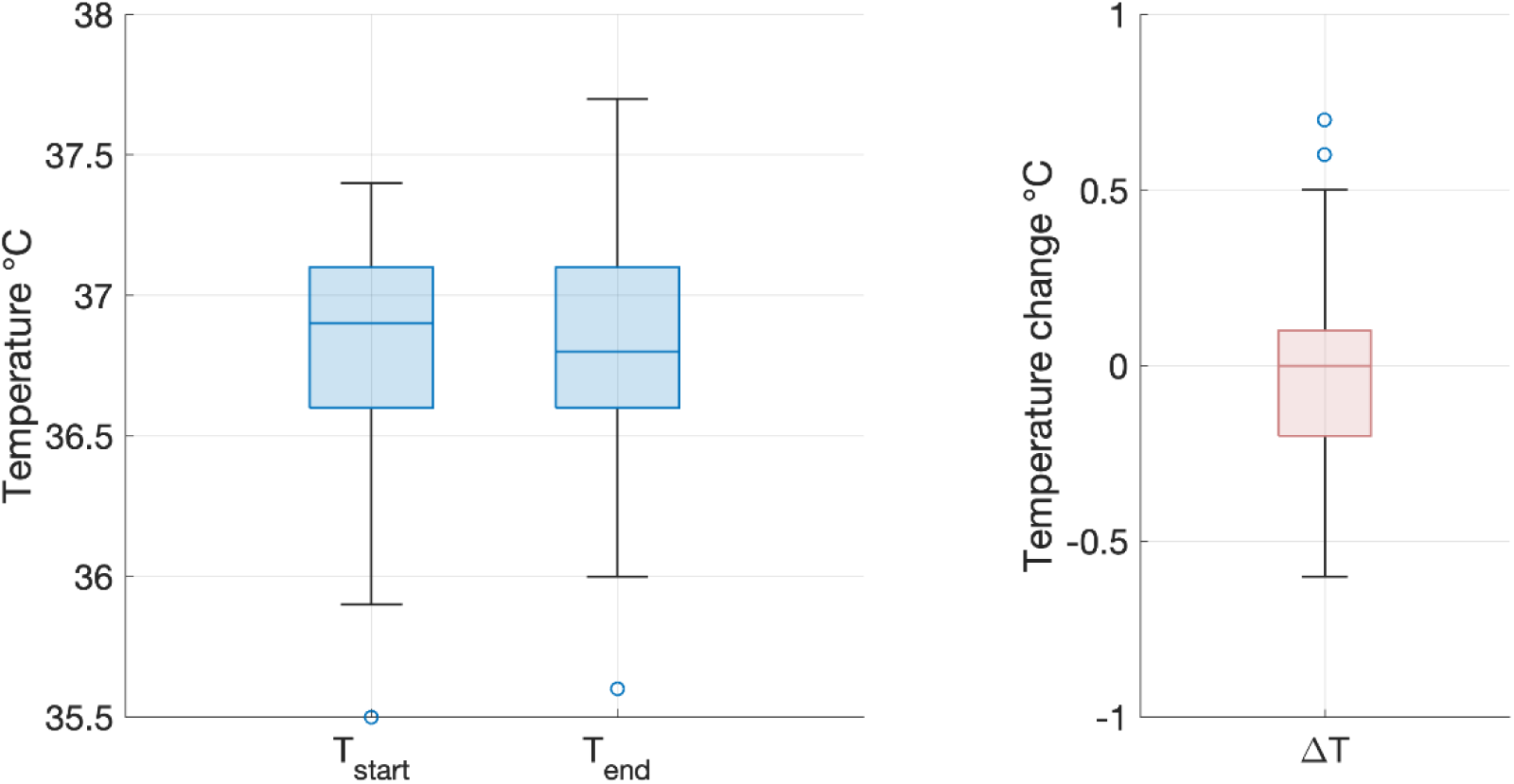
Infant temperature during scanning on the 7T system. There was no significant difference between infant axillary temperature at the start (*left*) and end (*right*) of the imaging session on the 7T system (paired two tailed t-test: p=0.76). Box and whisker plots showing data median (bold line), 25^th^ and 75^th^ centile (box borders) and data range (whiskers). Data outliers are denoted by circles.

Images from the AFI sequence (*Figure 3)* show that relative B_1_ ^+^ magnitude is non-uniform over the brain. The example map shown (Figure 3a) is typical of what was seen across all infants studied, with a ‘center brightening’ of the B_1_ ^+^ magnitude, and lower values seen peripherally, particularly in superior regions. The FSL brain extraction tool (BET) ^26^ was used to define a brain mask to then compute whole-brain histograms of B_1_ ^+^ magnitude, shown in Figure 3b. The histograms are consistent across all infants and show that the median B_1_ ^+^ over the brain was in the range 0.69-0.78 (relative to nominal value) with the central bright spot reaching values approximately 20-30% higher than nominal value.

**Figure 3:**
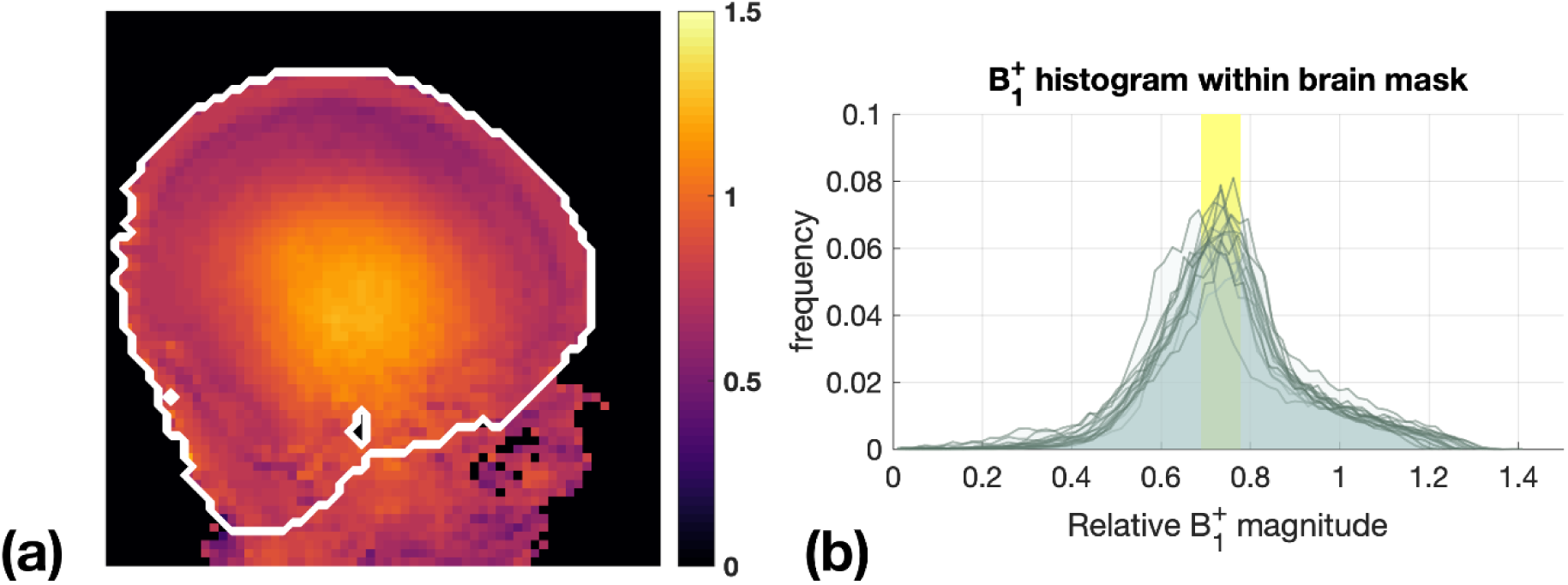
B_1_^+^ maps from 15 infants. (a) Example B_1_^+^ map obtained with AFI sequence, shown in relative units (i.e., relative to nominal B_1_^+^). The white contour marks the approximate brain outline. (b) Histograms of relative B_1_^+^ magnitude within the brain mask in 15 subjects. The yellow box shows the range of median relative B_1_^+^ (0.69-0.78).

In infants with paired studies, expert neuroradiology review of the T2 weighted anatomical images acquired at 7T was considered in all cases to be equivalent or higher in comparison to those acquired at 3T. In addition, the higher tissue contrast and spatial resolution of the 7T images resulted in improved visualization of specific structures which are typically challenging to delineate at lower field strengths (*Figure 4*). These included the hippocampi, greater definition of substructures within the deep grey matter structures (thalami and basal ganglia), and cortical folding in areas where it tends to be particularly convoluted and therefore difficult to delineate (i.e., the occipital lobe). Furthermore, additional detail which is not appreciated at lower field strengths could be seen, including the white matter medullary veins and cerebellar foliage.

**Figure 4:**
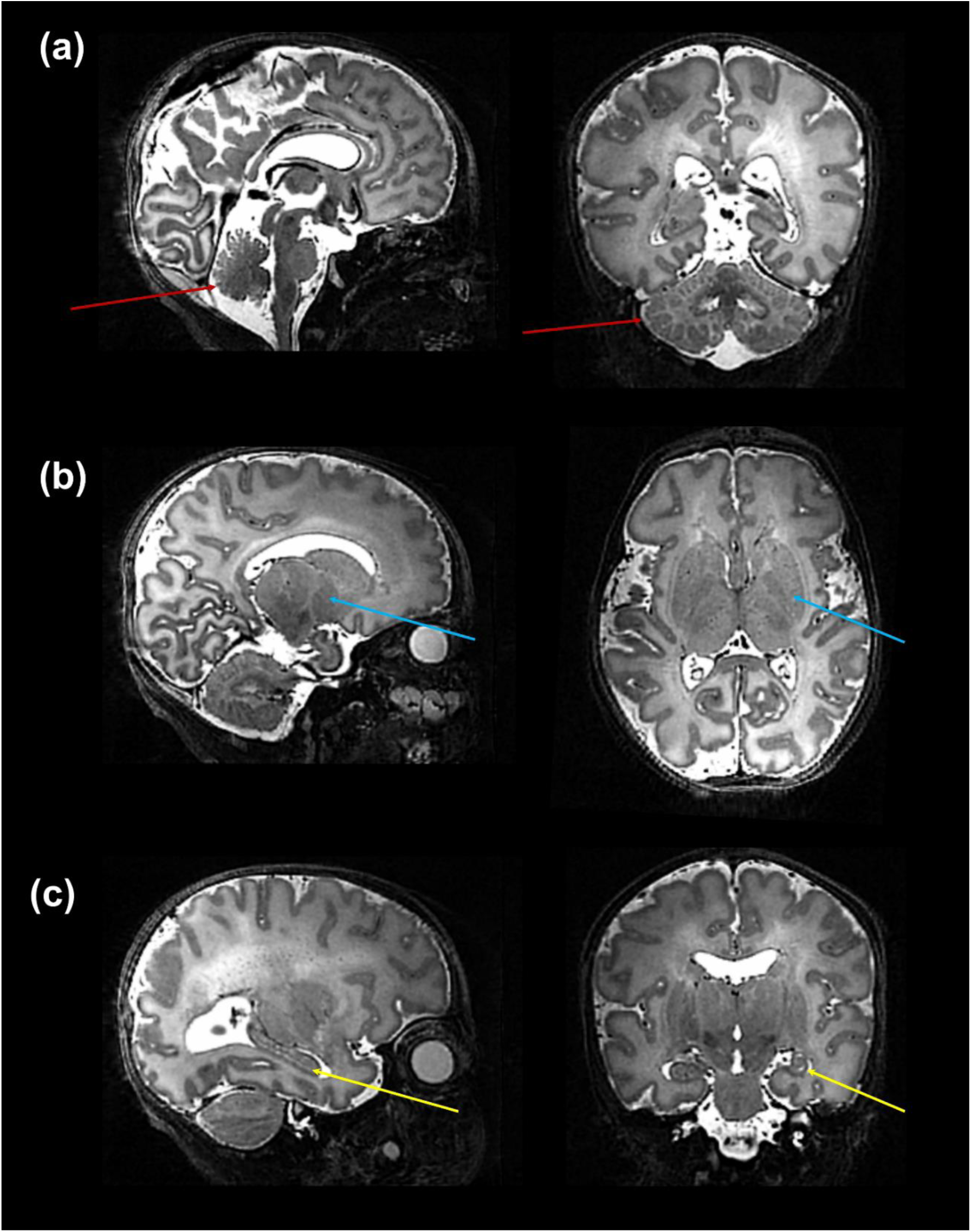
High resolution T2 weighted 7T image acquired from a preterm infant. Images show excellent visualisation of the: (a) cerebellar vermis and foliage (*red arrows*); (b) deep gray matter nuclei within the basal ganglia and thalamus (*blue arrows*); (c) hippocampus (*yellow arrow*).

Specific pathological features that could be clearly visualized on 7T images included cystic lesions and septi in periventricular leukomalacia (PVL), areas of micro-hemorrhage, subdural cerebral hemorrhage, cortical polymicrogyria, and absence of the cavum septum pellucidum (*Figure 5*). SVRTK reconstructed images further improved image contrast and quality by correcting motion and inter-slice artifacts (*Figures 6 a,b,c*), enabling robust automated tissue segmentation and surface generation (*Figure 6d*). Image quality of the native T2-weighted images was assessed to be good in 64% of the acquisitions (*Figure 1b*), which increased further to 82% following SVR reconstruction (*Figure 1c)*, which compares favorably against equivalent figures assessed with the same criteria on state-of-the-art images acquired for the dHCP with a 3T system and dedicated neonatal receive coil ^19^.

**Figure 5:**
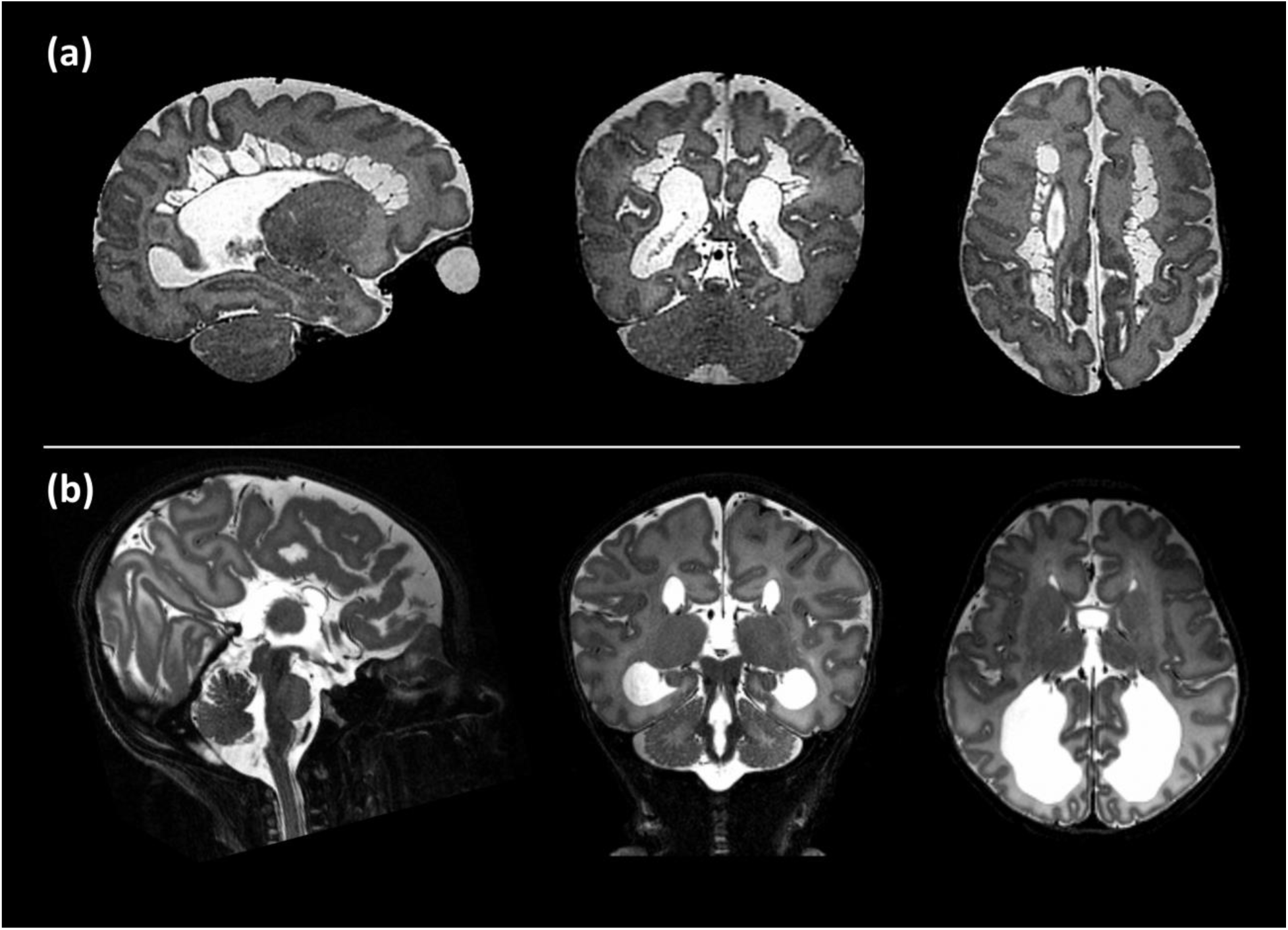
Examples of intracranial pathology identified on T2 weighted images at 7T. *(from left to right: illustrative slices in the sagittal, coronal, and axial planes).* Shown are: (a) Cystic PVL; (b) Complete agenesis of the corpus callosum.

**Figure 6:**
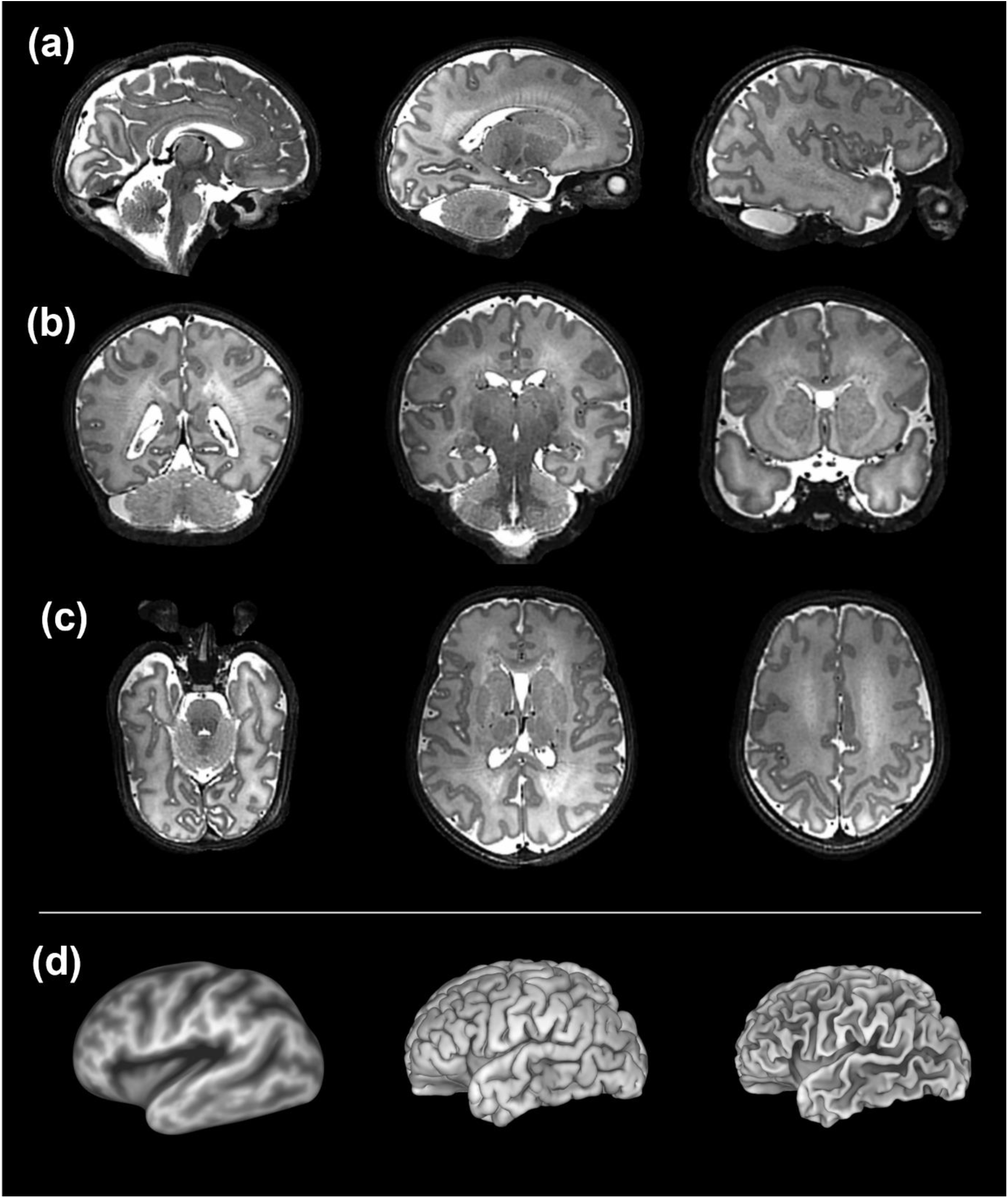
Slice to Volume reconstructed T2 weighted images acquired from a preterm infant shown in the (a) sagittal, (b) coronal and (c) axial planes. These high contrast, high resolution images are amenable for further processing such as the generation of (d) (*from left to right*) inflated, pial, and white matter surfaces.

The higher static magnetic field strength at 7T additionally led to marked gains in sensitivity in image contrasts dependent on magnetic susceptibility such as SWI. In addition to providing detailed visualization of the cerebral vasculature including both the arteries and veins (*Figure 7a*), the clinical value of SWI acquisitions was evident for identification of small areas of intracerebral hemorrhages in infants with congenital cardiac disease, in a preterm infant with a white matter cystic lesion where the hemorrhagic origin of the lesion could be appreciated (*Figure 7b*), and in a preterm infant with extensive intracerebral hemorrhage in the distribution of the deep medullary veins (*Figure 7c*). Gains in sensitivity and spatial specificity could also be appreciated using BOLD fMRI, with more cortically localized and broader repertoire of resting-state networks in comparison to 3T ^3^ (*Figure 8*). Expected gains in sensitivity and spectral resolution were also realised with MRS indicating the possibility of resolving spectral contribution of GABA, glutamate and glutamine that would usually require an edited acquisition scheme at lower field strengths (*Figure 9*).

**Figure 7:**
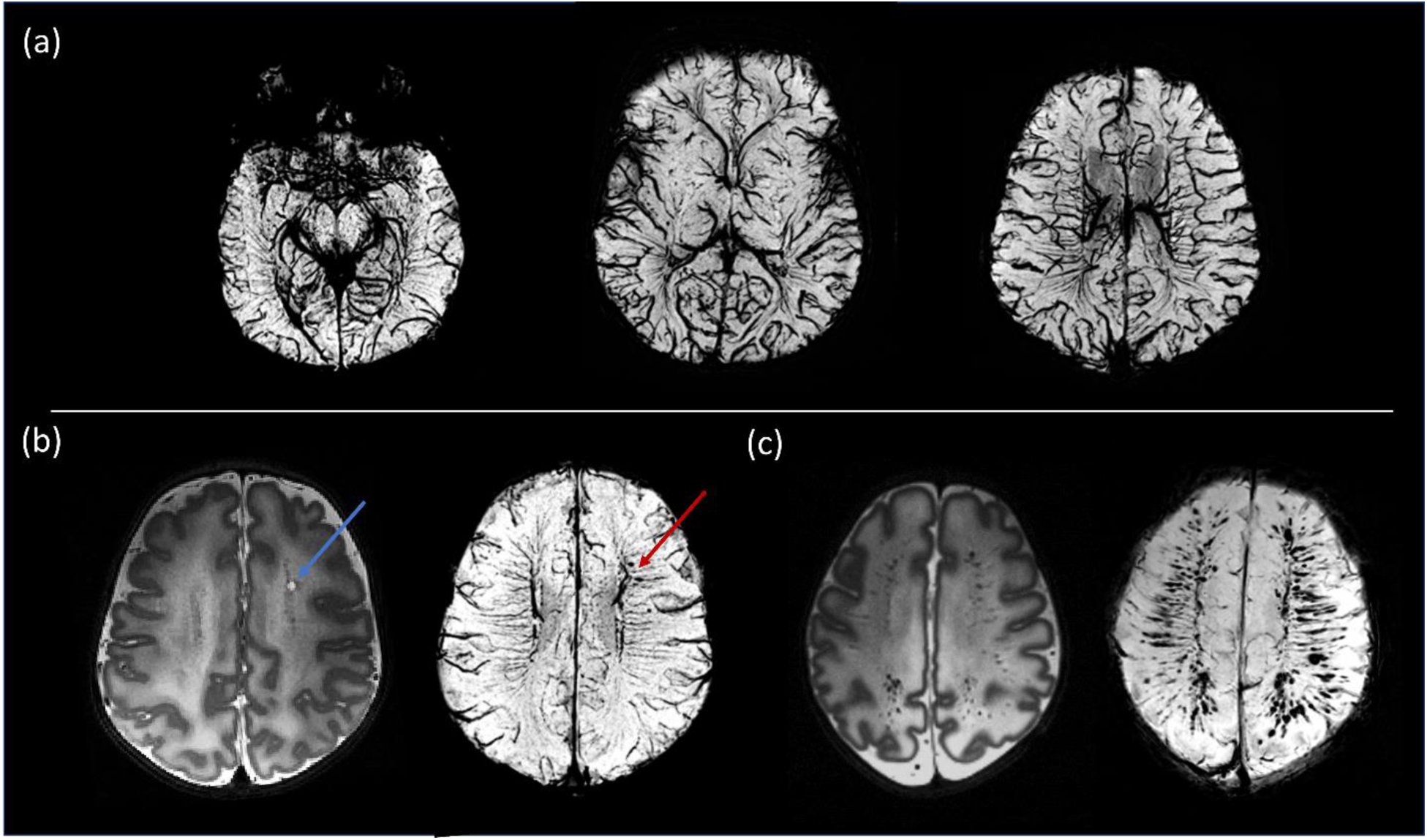
Susceptibility weighted images (SWI). (a) Example MIP axial slices from the SWI data acquired from a healthy neonate imaged at term equivalent age. (b) A cystic lesion (blue arrow) noted on T2- weighted images (*bottom left*) in a preterm infant. SWI demonstrated the hemorrhagic origin of the lesion (red arrow) and gives possible insight into the underlying pathophysiology through its adjacent location to the deep medullary veins. (c) Preterm infant with extensive intracerebral hemorrhage in the distribution of the medullary veins. The extent of this is visualized more clearly on the SWI image (bottom right).

**Figure 8:**
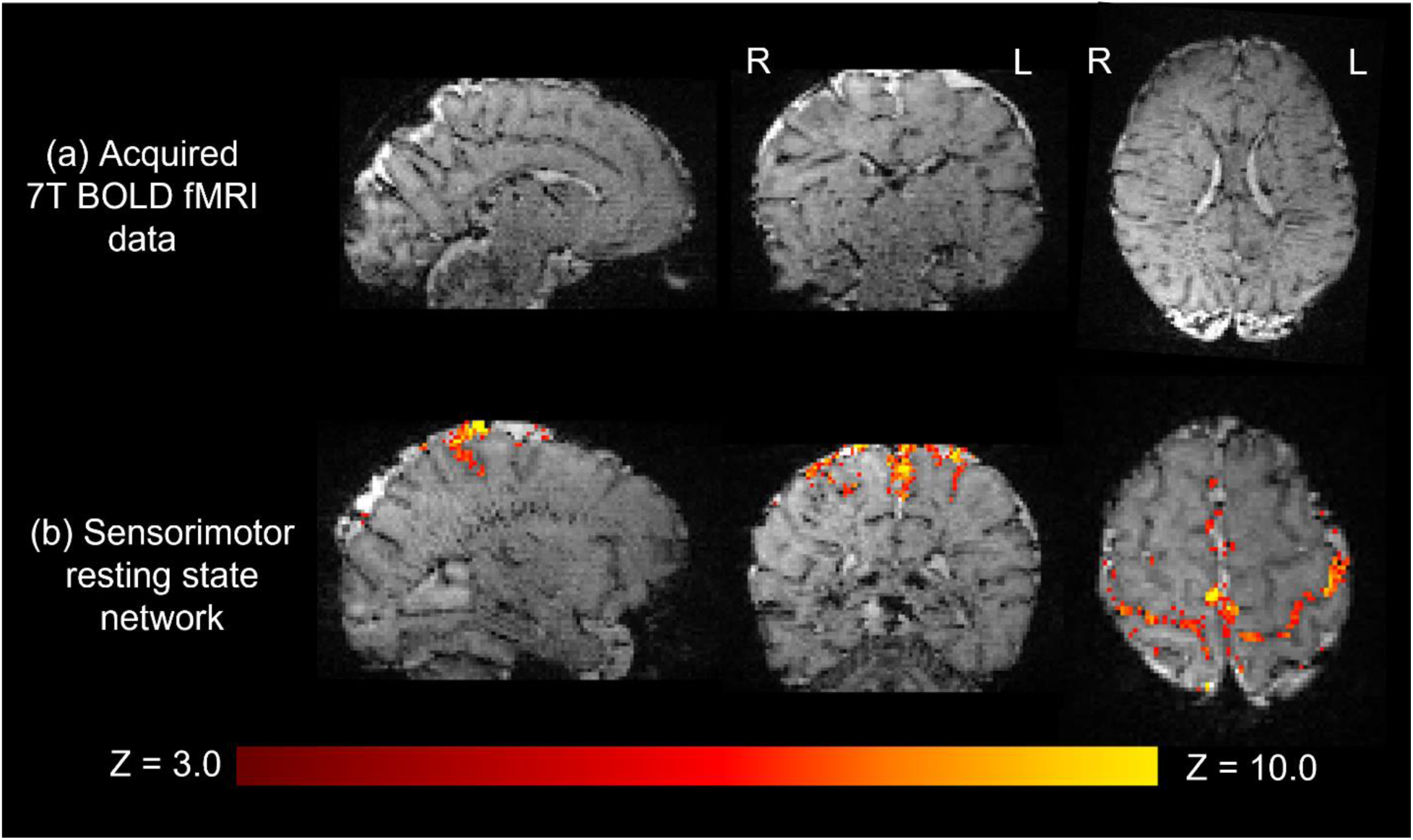
Resting state functional MRI data acquired from a preterm infant at 7T. (a) High spatial resolution (1mm isotropic) high contrast whole brain BOLD fMRI data acquired at 7T from a preterm infant. (b) The sensorimotor resting state network derived using independent component analysis. Activation can be seen to clearly localize to the cortical ribbon, following the configuration of the sulci.

**Figure 9:**
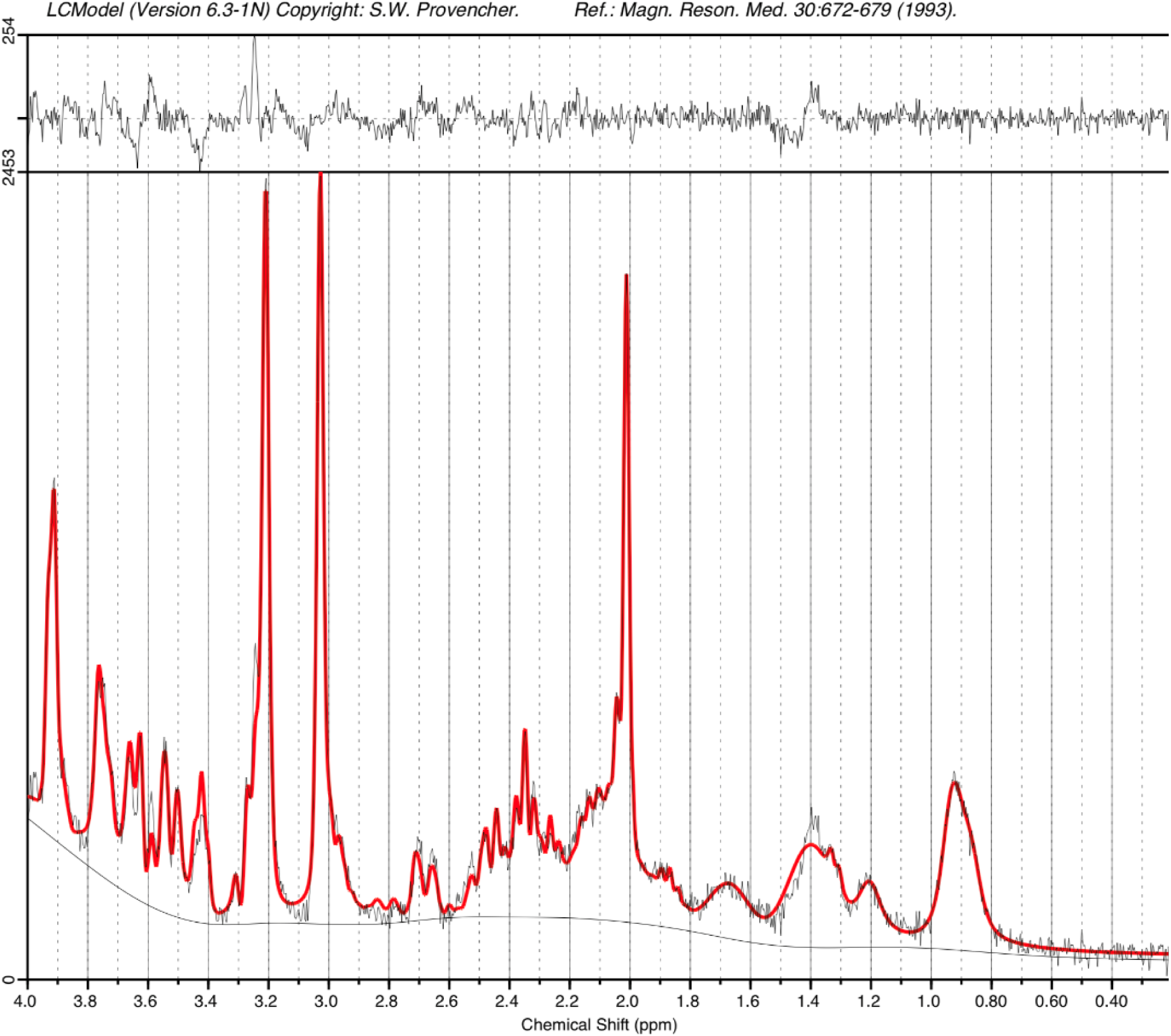
STEAM MRS spectrum acquired from an infant at term equivalent age on the 7T system. A narrow linewidth (4.8Hz) and high SNR (26) as estimated by LCModel are observed. The acquisition time was just 2’24”.

## DISCUSSION

We report our first experiences and demonstrate feasibility of imaging neonates on a 7T system and the potential gains in anatomical and pathological sensitivity in brain images. In all cases, infants tolerated image acquisition at ultra-high field with stable vital signs and body temperature throughout the examination. Acquired high resolution anatomical images demonstrated high tissue contrast enabling visualization of several structures which are typically challenging to delineate in neonates at standard field strengths. The potential of ultra-high field imaging to provide new insight about the developing neonatal brain was further highlighted using contrasts dependent on tissue magnetic susceptibility including SWI and fMRI; and for MRS data which benefits both from increased SNR and spectral dispersion.

Recent studies of electromagnetic and radiofrequency exposure for 7T imaging of neonates suggested that in contrast to imaging at standard field strengths, more conservative operational limits should be used to limit SAR at ultra-high field in comparison to adults ^11^. Addressing the potential risks of body temperature heating is further complicated in neonates by their under-developed thermal regulation and high surface-to-volume body ratio, which means that they can also become rapidly hypothermic if not appropriately insulated with clothes and/or blankets ^13, 27^. Accounting for these factors is imperative as not only can neonates not directly indicate if they are uncomfortable or feeling cold/warm in the scanner, but thermal instability can result in significant physiological stress including increased oxygen consumption, decreased cardiac output and metabolic acidosis ^28^. Here we demonstrate that neonatal body temperature remained stable during ultra-high field imaging, whilst using a combination of appropriate adaptation of scanner software models and practical steps including swaddling and continuous monitoring using MR compatible equipment.

In addition to informing our safety assessment, the prior modelling study ^11^ predicted that B_1_ ^+^ would be non-uniform for neonates using the 7T head coil, contrary to expectations that the small size of neonatal subjects might mitigate this problem. The measured B_1_ ^+^ distributions (Figure 3) from the current study are consistent with predictions from the aforementioned modelling study. Figure 5 in Clarke et al shows equivalent measurements from adults using the same coil ^29^; it is noticeable that while the centre-brightening effect is comparable between neonates and adults, areas of significant B_1_ ^+^ drop-out are less extreme in neonates than adults. These characteristics should be factored in when optimizing sequences for this cohort.

The clinical value of ultra-high field brain imaging has become relatively well established in older children and adults, particularly in applications and pathologies where the higher spatial resolution and improved tissue contrast has proven to increase sensitivity and offer new insight. Key examples include management of drug-resistant epilepsy where it can markedly improve detection and differentiation of potentially treatable epileptogenic lesions in comparison to imaging at standard field strengths, leading to international consensus recommendations for 7T imaging for this specific application ^30^; improved delineation of vascular pathology such as aneurysms or arteriovenous malformations ^31^; detailed visualization of the deep grey matter structures which can help guide the insertion of deep brain stimulation electrodes ^32^; and detection and differentiation of lesions in multiple sclerosis ^33^.

Whilst the clinical indications for 7T imaging of neonates are yet to be established, our initial experiences similarly suggest that there is clear potential through improving sensitivity for conditions which impact the developing cortex, vasculature, and smaller structures. This includes following preterm birth, which globally now represents greater than 15 million births annually and has significant implications for later neurodevelopmental outcome in survivors ^34^. In addition to the traditionally recognized patterns of direct white matter injury such as cystic PVL, preterm birth is also associated with subtle abnormalities such as punctate white matter lesions or micro-hemorrhages for which the underlying pathophysiology and clinical meaning remain uncertain ^35^. Furthermore, it is increasingly appreciated that adverse neurocognitive outcomes following preterm birth are related to lifelong alterations in cortical development, thalamic growth and connectivity ^36^. Infants with congenital cardiac disease are at high risk of peri-operative hemorrhage or ischemic injury, and often have chronically reduced cerebral oxygenation^37^. These factors have also been found to result in profound life-long effects on cortical maturation which are similarly associated with adverse neurodevelopmental outcomes ^38^. Thus, imaging neonates at 7T could not only enhance detection of injuries in these high-risk populations, but also holds clear potential to provide marked new mechanistic insight into the pathophysiology underlying their neurodevelopmental consequences.

We also highlight the potential of gaining new insights into the developing brain using specific image contrasts which benefit from the increased field strength such as SWI, MRS and fMRI. The SWI method in particular has been shown to have particular value at 7T, where it has broad clinical applications including improved characterization of gliomas for surgical planning and epileptogenic foci ^30^, but additionally has potential to provide important new insights into the pathophysiology of common conditions such as dementia ^39^. Here, we show that SWI in neonates not only has high sensitivity for detecting areas of hemorrhage, but additionally can provide detailed visualization of the cerebral vasculature including both the arterial and venous systems (*see Figure 7*). This knowledge could provide novel insight about the specific role of the vascular system and cerebral perfusion in regional brain development and how this relationship is altered by pathology. fMRI BOLD contrast is similarly enhanced at ultra-high field and has been shown to enable fine-scale studies of the brain’s functional architecture at higher resolution and sensitivity compared to standard field strengths, including delineation of activity within cortical columns, layers and specific deep grey matter nuclei ^8^. The ability to characterize brain activity at this level of detail in the neonatal brain is a compelling prospect, as this is a crucial time for the establishment of the brain’s life-long framework of functional connectivity, with studies at 3T demonstrating rapid maturation of resting state networks in the time leading up to birth ^3^. Importantly, early-life alterations in functional connectivity such as those associated with preterm birth persist into later life and correlate with adverse neurodevelopmental outcome ^4, 36^. Significantly increased sensitivity and spectral resolution at 7T with 1H-MRS have been well described in adults ^9^, which we were also able to demonstrate in neonates. Wider application could potentially inform about how inhibitory-excitatory neurotransmitter balance evolves across early development and about its possible role in the pathophysiology of neurodevelopmental disorders such as autism ^4^.

We describe our initial experiences and demonstrate feasibility of 7T neonatal imaging with a limited range of sequences optimized for this population. Although the acquired images had high SNR, an important consideration for this work is that we used a standard adult transmit and receive head coil. Previous work at 3T has demonstrated that SNR, image homogeneity and head immobilization can all be considerably improved with a head coil which is appropriately sized so that the receive elements are closer to the neonatal head ^17^. Work is thus underway to develop a neonatal specific head coil for 7T scanning in our center ^40^. Whilst we demonstrate significant gains in anatomical detail and sensitivity with high-resolution T2-weighted imaging, high contrast T1-weighted imaging was found to be challenging to acquire at 7T without knowledge of neonatal-specific tissue T1 values. Systematic data collection is therefore underway to establish brain tissue T1 and T2 relaxation values across the perinatal period in both preterm and term born neonates and will be reported separately. Nevertheless, our work highlights the clear potential of ultra-high field brain imaging in neonates to improve diagnosis and understanding of pathological mechanisms during this key stage of life. This has important implications not only for the clinical management of conditions known to originate in this period (such as cerebral palsy) but also for other common but hitherto poorly understood conditions like autism and mental health disorders which likely have their origin in the perinatal period.

## Data Availability

All data produced in the present study are available upon reasonable request to the authors

## ACKNOWLEDGEMENTS

The work was supported a project grant awarded by Action Medical Research [GN2728]. TA is also supported by an MRC Clinician Scientist Fellowship [MR/P008712/1] and Transition Support Award [MR/V036874/1]. PC, AT, JOM, MAR, ADE and TA received support from the Medical Research Council Centre for Neurodevelopmental Disorders, King’s College London [MR/N026063/1]. LCG received support by project PID2021-129022OA-I00, funded by MCIN / AEI / 10.13039/501100011033 / FEDER, EU. The authors also acknowledge support in part from the Wellcome Engineering and Physical Sciences Research Council (EPSRC) Centre for Medical Engineering at Kings College London [WT 203148/Z/16/Z]. The authors are grateful for discussion and advice from Jonathan Polimeni, David Carmichael, Essa Yacoub and colleagues at Utrecht University. We acknowledge significant input and support from colleagues at Siemens Healthineers without whom this work would not have been possible. We are also grateful for assistance from Philips Invivo, and Pearltec. This research was funded/supported by the National Institute for Health and Care Research (NIHR) Clinical Research Facility at Guy’s and St Thomas’ NHS Foundation Trust. The views expressed are those of the author(s) and not necessarily those of the NHS, the NIHR or the Department of Health and Social Care. The funders had no role in the design and conduct of the study; collection, management, analysis, and interpretation of the data; preparation, review, or approval of the manuscript; and decision to submit the manuscript for publication.

## REFERENCES

1. Kostovic I, Jovanov-Milosevic N. The development of cerebral connections during the first 20-45 weeks’ gestation. Semin Fetal Neonatal Med 2006;11:415–422.

2. Ben-Ari Y, Gaiarsa JL, Tyzio R, Khazipov R. GABA: a pioneer transmitter that excites immature neurons and generates primitive oscillations. Physiol Rev 2007;87:1215-1284.

3. Doria V, Beckmann CF, Arichi T, et al. Emergence of resting state networks in the preterm human brain. Proc Natl Acad Sci U S A 2010;107:20015–20020.

4. Batalle D, Edwards AD, O’Muircheartaigh J. Annual Research Review: Not just a small adult brain: understanding later neurodevelopment through imaging the neonatal brain. J Child Psychol Psychiatry 2018;59:350–371.

5. Pohmann R, Speck O, Scheffler K. Signal-to-noise ratio and MR tissue parameters in human brain imaging at 3, 7, and 9.4 tesla using current receive coil arrays. Magn Reson Med 2016;75:801–809.

6. van Lanen R, Colon AJ, Wiggins CJ, et al. Ultra-high field magnetic resonance imaging in human epilepsy: A systematic review. Neuroimage Clin 2021;30:102602.

7. Geurts LJ, Zwanenburg JJM, Klijn CJM, Luijten PR, Biessels GJ. Higher Pulsatility in Cerebral Perforating Arteries in Patients With Small Vessel Disease Related Stroke, a 7T MRI Study. Stroke 2018;50:STROKEAHA118022516.

8. Dumoulin SO, Fracasso A, van der Zwaag W, Siero JCW, Petridou N. Ultra-high field MRI: Advancing systems neuroscience towards mesoscopic human brain function. Neuroimage 2018;168:345–357.

9. Tkac I, Oz G, Adriany G, Ugurbil K, Gruetter R. In vivo 1H NMR spectroscopy of the human brain at high magnetic fields: metabolite quantification at 4T vs. 7T. Magn Reson Med 2009;62:868–879.

10. Annink KV, van der Aa NE, Dudink J, et al. Introduction of Ultra-High-Field MR Imaging in Infants: Preparations and Feasibility. AJNR Am J Neuroradiol 2020;41:1532–1537.

11. Malik SJ, Hand JW, Satnarine R, Price AN, Hajnal JV. Specific absorption rate and temperature in neonate models resulting from exposure to a 7T head coil. Magn Reson Med 2021;86:1299–1313.

12. Williams LA, Gelman N, Picot PA, et al. Neonatal brain: regional variability of in vivo MR imaging relaxation rates at 3.0 T--initial experience. Radiology 2005;235:595–603.

13. Plaisier A, Raets MM, van der Starre C, et al. Safety of routine early MRI in preterm infants. Pediatr Radiol 2012;42:1205–1211.

14. Malik SJ, Beqiri A, Price AN, Teixeira JN, Hand JW, Hajnal JV. Specific absorption rate in neonates undergoing magnetic resonance procedures at 1.5 T and 3 T. NMR Biomed 2015;28:344–352.

15. Bridgen P, Malik S, Wilkinson T, et al. Reliability and safety of anaesthetic equipment around an high-field 7-Tesla MRI scanner. Br J Anaesth 2023;130:e490–e492.

16. Yarnykh VL. Actual flip-angle imaging in the pulsed steady state: a method for rapid three-dimensional mapping of the transmitted radiofrequency field. Magn Reson Med 2007;57:192–200.

17. Hughes EJ, Winchman T, Padormo F, et al. A dedicated neonatal brain imaging system. Magn Reson Med 2017;78:794–804.

18. Kuklisova-Murgasova M, Quaghebeur G, Rutherford MA, Hajnal JV, Schnabel JA. Reconstruction of fetal brain MRI with intensity matching and complete outlier removal. Med Image Anal 2012;16:1550–1564.

19. Makropoulos A, Robinson EC, Schuh A, et al. The developing human connectome project: A minimal processing pipeline for neonatal cortical surface reconstruction. Neuroimage 2018;173:88–112.

20. Smith SM, Jenkinson M, Woolrich MW, et al. Advances in functional and structural MR image analysis and implementation as FSL. Neuroimage 2004;23 Suppl 1:S208–219.

21. Eckstein K, Bachrata B, Hangel G, et al. Improved susceptibility weighted imaging at ultra-high field using bipolar multi-echo acquisition and optimized image processing: CLEAR-SWI. Neuroimage 2021;237:118175.

22. Rodgers CT, Robson MD. Coil combination for receive array spectroscopy: Are data-driven methods superior to methods using computed field maps? Magn Reson Med 2016;75:473–487.

23. Near J, Harris AD, Juchem C, et al. Preprocessing, analysis and quantification in single-voxel magnetic resonance spectroscopy: experts’ consensus recommendations. NMR Biomed 2021;34:e4257.

24. Provencher SW. Estimation of metabolite concentrations from localized in vivo proton NMR spectra. Magn Reson Med 1993;30:672–679.

25. Wilson M, Reynolds G, Kauppinen RA, Arvanitis TN, Peet AC. A constrained least-squares approach to the automated quantitation of in vivo (1)H magnetic resonance spectroscopy data. Magn Reson Med 2011;65:1–12.

26. Smith SM. Fast robust automated brain extraction. Hum Brain Mapp 2002;17:143–155.

27. Soll RF. Heat loss prevention in neonates. J Perinatol 2008;28 Suppl 1:S57–59.

28. Wood T, Thoresen M. Physiological responses to hypothermia. Semin Fetal Neonatal Med 2015;20:87–96.

29. Clarke WT, Mougin O, Driver ID, et al. Multi-site harmonization of 7 tesla MRI neuroimaging protocols. Neuroimage 2020;206:116335.

30. Opheim G, van der Kolk A, Markenroth Bloch K, et al. 7T Epilepsy Task Force Consensus Recommendations on the Use of 7T MRI in Clinical Practice. Neurology 2021;96:327–341.

31. Radojewski P, Slotboom J, Joseph A, Wiest R, Mordasini P. Clinical Implementation of 7T MRI for the Identification of Incidental Intracranial Aneurysms versus Anatomic Variants. AJNR Am J Neuroradiol 2021;42:2172–2174.

32. Xiao Y, Zitella LM, Duchin Y, et al. Multimodal 7T Imaging of Thalamic Nuclei for Preclinical Deep Brain Stimulation Applications. Front Neurosci 2016;10:264.

33. Kolb H, Absinta M, Beck ES, et al. 7T MRI Differentiates Remyelinated from Demyelinated Multiple Sclerosis Lesions. Ann Neurol 2021;90:612–626.

34. Cao G, Liu J, Liu M. Global, Regional, and National Incidence and Mortality of Neonatal Preterm Birth, 1990-2019. JAMA Pediatr 2022;176:787–796.

35. de Bruijn CAM, Di Michele S, Tataranno ML, et al. Neurodevelopmental consequences of preterm punctate white matter lesions: a systematic review. Pediatr Res 2022.

36. Rogers CE, Lean RE, Wheelock MD, Smyser CD. Aberrant structural and functional connectivity and neurodevelopmental impairment in preterm children. J Neurodev Disord 2018;10:38.

37. Bonthrone AF, Stegeman R, Feldmann M, et al. Risk Factors for Perioperative Brain Lesions in Infants With Congenital Heart Disease: A European Collaboration. Stroke 2022;53:3652–3661.

38. Kelly CJ, Christiaens D, Batalle D, et al. Abnormal Microstructural Development of the Cerebral Cortex in Neonates With Congenital Heart Disease Is Associated With Impaired Cerebral Oxygen Delivery. J Am Heart Assoc 2019;8:e009893.

39. van Rooden S, Versluis MJ, Liem MK, et al. Cortical phase changes in Alzheimer’s disease at 7T MRI: a novel imaging marker. Alzheimers Dement 2014;10:e19–26.

40. Clement J, Tomi-Tricot R, Malik SJ, Webb A, Hajnal JV, Ipek O. Towards an integrated neonatal brain and cardiac examination capability at 7 T: electromagnetic field simulations and early phantom experiments using an 8-channel dipole array. MAGMA 2022;35:765–778.

